# New *ZNHIT3* Variants Disrupting snoRNP Assembly Cause Prenatal PEHO Syndrome with Isolated Hydrops

**DOI:** 10.1101/2024.08.26.24312490

**Authors:** Md Lutfur Rahman, Adeline A. Bonnard, Feng Wang, Lyse Ruaud, Fabien Guimiot, Yangping Li, Ines Defer, Yilin Wang, Virginie Marchand, Yuri Motorin, Bing Yao, Séverine Drunat, Homa Ghalei

**Affiliations:** Department of Biochemistry, Emory University School of Medicine, Atlanta, Georgia, USA; Département de Génétique, Hôpital Robert Debré, Assistance Publique des Hôpitaux de Paris (AP-HP), Paris, France; INSERM UMR 1131, Saint-Louis Research Institute, Paris University, Paris, France; Department of Human Genetics, Emory University School of Medicine, Atlanta, Georgia, USA; INSERM UMR 1141, Paris-Cité University, NeuroDiderot, Paris, France; Université de Lorraine, SMP IBSLor, Biopôle, 9 Avenue de la Forêt de Haye, Vandoeuvre-les-Nancy, France; Université de Lorraine, UMR7365 IMoPA, CNRS, Biopôle, 9 Avenue de la Forêt de Haye, Vandoeuvre-les-Nancy, France

**Keywords:** Intrauterine fetal loss, fetal hydrops, developmental disorder, PEHO syndrome, snoRNP biogenesis, translation

## Abstract

ZNHIT3 (zinc finger HIT type containing protein 3) is an evolutionarily conserved protein required for ribosome biogenesis by mediating the assembly of small nucleolar RNAs (snoRNAs) of class C/D into ribonucleoprotein complexes (snoRNPs). Missense mutations in the gene encoding ZNHIT3 protein have been previously reported to cause PEHO syndrome, a severe neurodevelopmental disorder typically presenting after birth. We discuss here the case of two fetuses from a single family who presented with isolated hydrops during the early second trimester of pregnancy, resulting in intrauterine demise. Autopsy revealed no associated malformation. Through whole-genome quartet analysis, we identified two novel variants within the *ZNHIT3* gene, both inherited from healthy parents and occurring as compound heterozygotes in both fetuses. The c.40T>C p.Cys14Arg variant originated from the father, while the c.251_254delAAGA variant was of maternal origin. Analysis of the variants in human cell culture models reveals that both variants reduce cell growth, albeit to different extents, and impact the protein’s stability and function in distinct ways. The c.251_254delAAGA results in production of a stable form of ZNHIT3 that lacks a domain required for mediating snoRNP biogenesis, whereas the c.40T>C p.Cys14Arg variation behaves similarly to the previously described PEHO-associated *ZNHIT3* variants that destabilize the protein. Interestingly, both variations lead to a marked decrease in specific box C/D snoRNA levels, reduced rRNA levels and cellular translation. Analysis of rRNA methylation pattern in fetus samples reveals distinct sites of hypo 2’-O-methylation. RNA-seq analysis of undifferentiated and differentiated SHSY5Y cells transfected with the *ZNHIT3* variants reveals differential expression of a set of genes, many of which are associated with developmental processes and RNA binding compared to cells expressing wild-type ZNHIT3. In summary, this work extends the phenotype of PEHO syndrome to include antenatal manifestations and describe the molecular defects induced by two novel *ZNHIT3* variants.

## Introduction

Protein homeostasis regulation is crucial for cell survival and development in all living organisms. Therefore, the production of cellular ribosomes, responsible for protein synthesis, is tightly regulated by the action of many assembly factors, including proteins and noncoding RNAs.^1-4^ A critical step in ribosome biogenesis involves the synthesis, processing, and chemical modification of ribosomal RNAs (rRNAs).^5-7^ Highly abundant small nucleolar RNAs (snoRNAs) are required for rRNA processing and guiding the majority of rRNA chemical modifications in eukaryotes.^8^ Dysregulation of snoRNA levels or their function is associated with several human diseases ranging from cancer to immunodeficiency, metabolic disorders and neurological disorders.^9-16^ snoRNAs are categorized into two primary types based on their conserved sequence motifs. SNORDs, classified by their conserved C/D and C’/D’ sequence motifs, primarily guide 2’-O-methylation although two of them are involved in guiding rRNA acetylation.^17^ The second major group, SNORAs, characterized by their conserved box H/ACA elements, guide the conversion of uridines to pseudouridines.^18^ Besides those involved in chemical modifications, several snoRNAs are essential for rRNA processing and cleavage, and a subset of them has no known targets or functions and is therefore considered orphans.^19^

snoRNAs bind to a set of evolutionarily conserved core proteins to form active ribonucleoprotein complexes (snoRNPs) catalyzing rRNA modification. Box C/D snoRNPs consist of a set of four essential core proteins, including SNU13, NOP56 and NOP58, and the 2′-O-methyltransferase FBL (fibrillarin).^20,21^ Formation of box C/D snoRNP complexes requires the action of assembly factors that are conserved from yeast to man. ZNHIT3 (human zinc finger HIT type containing protein 3), is a nucleolar protein critical for assembly of box C/D snoRNPs. ZNHIT3 participates in a network of protein-protein interactions including NUFIP1 protein.^22,23^ Studies using human cell lines and yeast have demonstrated that ZNHIT3 (Hit1 in yeast) is required for maintaining the steady-state levels of NUFIP1 (Rsa1 in yeast).^22^ Systematic quantitative stable isotope labeling by amino acids in cell culture proteomic experiments revealed that ZNHIT3 is present in a protein-only complex required for snoRNP assembly and is released upon binding of C/D snoRNAs.^24^ Deletion of the *HIT1* gene in yeast leads to reduced steady-state levels of box C/D snoRNAs and hinders the assembly of box C/D snoRNPs as well as ribosome synthesis.^22^ The NUFIP1-ZNHIT3 complex is shown to aggregate on autophagosomes and lysosomes following the inhibition of mTORC1, triggering ribophagy.^25^

Pathogenic variants in the *ZNHIT3* gene have been identified as the underlying cause of a severe neurodevelopmental condition known as PEHO syndrome. This syndrome is characterized by postnatal progressive cerebellar atrophy, edema, hypsarrhythmia, optic atrophy, arrest of psychomotor development, and a poor prognosis.^26-28^ Only two PEHO-linked missense *ZNHIT3* variants have been described so far, which lie within the Zf-HIT domain of the protein. The founder Ser31Leu (S31L) variant was consistently identified as the cause of autosomal recessive PEHO syndrome in a cohort of Finnish descent. The second variant, Cys14Phe (C14F), was identified in one patient in the compound heterozygous state along with the S31L variant.^28,29^ Studies in human cell cultures have shown that the ZNHIT3-S31L variation destabilizes the protein, decreasing steady-state protein levels.^28^ Studies of these protein variants modeled in budding yeast demonstrated that missense mutations identified in individuals with PEHO syndrome reduce the steady-state levels of Hit1 protein in yeast. This decrease in protein levels is accompanied by a notable reduction in the cellular amounts of box C/D snoRNAs, site-specific rRNA modification changes, and impairment of rRNA processing and cellular translation.^30^

Here, we report for the first time a fetal form of PEHO syndrome caused by two novel *ZNHIT3* variants in the compound heterozygous state. We provide functional studies showing that the new variants affect gene expression, snoRNP biogenesis, snoRNA levels, rRNA modification, and translation.

## Materials and methods

### Patient and clinical information collection

Complete autopsies of both fetuses were performed with the informed consent of both parents, in accordance with French regulations and according to standardized protocols including radiographs, photographs, macroscopic and histological examinations of all viscera. Fetal biometric data were assessed by first-trimester sonography crown-rump length measurement and confirmed at autopsy by the evaluation of the fetal biometry, and organ and skeletal maturations as described previously.^31^ Tissue samples from the lungs were collected during the autopsy for cryopreservation and potential future extraction of DNA or RNA.

Brains were removed and fixed in 10% buffered formalin added with 3 g/L of ZnSO_4_ for approximately 2 to 3 weeks. Brains were then cut into coronal sections of 0.5 cm of thickness for routine examination. Seven-micrometer-thick paraffin-embedded tissue sections were performed on Superfrost Plus slides (Thermo Scientific) and stained with hematoxylin and eosin (H&E). Brains of fetuses resulting from late miscarriages without cerebral anomalies at autopsy were selected as controls.

### Molecular analysis

Whole genome sequencing (WGS) analysis was performed in quartet on DNA extracted from cryopreserved fetal lung, and peripheral blood samples from parents with their informed consent.

### HEK293T cell culture and reagents

Human embryonic Kidney (HEK293T) cells were obtained from ATCC and confirmed to be negative for mycoplasma using the Southern Biotech Mycoplasma Detection Kit. HEK293T cells were propagated in Dulbecco’s modified Eagle’s medium (D-MEM; Corning) supplemented with 10% fetal bovine serum (FBS; Corning) with 1% penicillin-streptomycin (Gibco). The cells were maintained at 37°C with 5% CO_2_ in a humidified incubator. For transfection, HEK293T cells were seeded in 6-well tissue culture plates at a density of 2x10^5^ cells/well and transfected with plasmids (2 µg total plasmid per well) on the following day using the Lipofectamine 3000 (Thermo Fisher Scientific) or PEI MAX reagent (Polysciences).

### Lymphoblastoid cell lines

Lymphoblastoid cell lines from parents were maintained in RPMI 1640 medium (Deutscher) supplemented with 10% fetal calf serum (Gibco), 1% penicillin-streptomycin (Gibco), 2 mM glutamine, and 1% HEPES (Eurobio Scientific). RNA from these cells was used for RiboMethSeq analysis.

### Immunohistochemical staining

ZNHIT3 protein localization and expression in formalin-fixed paraffin-embedded (FFPE) brain tissues were performed with the OptiView DAB IHC Detection Kit (Ventana Medical Systems) on a BenchMark GX automated system (Ventana Medical Systems) using a rabbit anti-ZNHIT3 primary monoclonal antibody (1:50; HPA078211, Sigma Aldrich). Signal intensity was enhanced using the OptiView Amplification Kit. Slides were digitized, and immunostaining intensities were compared using the ImageJ software.

### Generation of ZNHIT3 variant plasmids

The HA-tagged pRK5-ZNHIT3 plasmid (Addgene #110953) was modified by site-directed mutagenesis to construct four plasmids expressing HA-tagged ZNHIT3 variants (ZNHIT3-C14R, ZNHIT3-C14F, ZNHIT3-S31L and ZNHIT3-Δ251-254). The Lentiviral plasmids of ZNHIT3-WT, ZNHIT3-C14R, and ZNHIT3-Δ251-254 were generated from the FUGW plasmid (Addgene #14883). All plasmids were validated by Sanger sequencing.

### Immunoblotting and assay of protein stability

HEK293T cells were transfected with plasmids expressing HA-tagged wild type or variant ZNHIT3. All plasmids used in this study are listed in **Table S1**. The cells were washed 48 h post-transfection with phosphate-buffered saline (PBS) and lysed in 250 µL of RIPA lysis buffer [150 mM sodium chloride, 20 mM Tris-HCl pH 8.0, 1% NP-40, 10% sodium deoxycholate, 10% SDS]. Total cell lysates were cleared by centrifugation (13000 rpm, 15 min, 4°C), and the total protein concentration was measured using a BCA kit (Pierce). Equal total protein concentrations were analyzed by SDS-PAGE, followed by Immunoblotting and quantification using Image Lab (Bio-Rad). To evaluate the stability of the ZNHIT3 variants compared to the wild-type protein, transfected HEK-293 cells were treated with 10 μg/mL cycloheximide in DMEM at 37°C. Cells were harvested at time 0 and after 2, 4, and 6 hours of incubation with cycloheximide, washed with ice-cold PBS and lysed in RIPA buffer. ZNHIT3 levels were measured by immunoblotting using an anti-HA antibody.

### Immunoprecipitation

For immunoprecipitation assays, transfected HEK293T cells were washed with ice-cold PBS and lysed in cold RIPA buffer containing protease inhibitors (Complete Mini, EDTA-free; Roche). Lysates were kept on ice and clarified by centrifugation in a microcentrifuge at 13,000 rpm at 4°C for 10 min. The clarified lysate was incubated with anti-HA-tag magnetic beads with gentle rotation at 4°C for 90 min. The resin was washed three times with ice-cold lysis buffer. 0.1 M glycine at pH 2.5 was used for eluting proteins. Immunoprecipitated proteins were denatured by the addition of SDS loading buffer and boiling for 5 min. The eluted proteins were analyzed on SDS-PAGE followed by immunoblotting.

### Lentivirus production and infection

Viruses were produced using FUGW-ZNHIT3 plasmids (ZNHIT3-WT, ZNHIT3-C14R, and ZNHIT3-Δ251-254) transfected into HEK 293T/17 cells (ATCC) with polyethyleneimine (PEI 25K, Polyscience). In brief, cells were seeded at 3.8×10^6^ cells in a 10 cm tissue culture plates and incubated overnight at 5% CO_2_ at 37 °C. 1.3 pmol of psPAX2 (Addgene, #12260), 0.72 pmol of pMD2.G (Addgene, #12259), and 1.64 pmol of FUGW-ZNHIT3 plasmid were diluted in 500 μL Opti DMEM, then mixed with 500 μL DMEM with PEI (ratio 1 μg DNA: 3 μg PEI at 1 mg/mL). Virus within the complete medium was harvested at 48, 72 hours post-transfection and centrifuged at 500 xg for 5 minutes to remove the packaging cells. The supernatant was filtered using a 0.45 μm PES filter and then concentrated with a Lenti-X concentrator (TaKaRa, #631231). The prepared virus was aliquoted and stored at -80°C. The produced virus was validated through infection of HEK293T cells followed by immunoblotting to measure the ZNHIT3 protein level using the signal from the HA tag. The undifferentiated-infected SHSY-5Y cells were collected 48 hours post-infection, and the differentiated-infected cells were collected 18 days post-infection.

### SHSY-5Y cell culture and differentiation

SH-SY5Y neuroblastoma cells were obtained from ATCC and confirmed negative for mycoplasma using the Southern Biotech Mycoplasma Detection Kit. Cells were maintained in 15% fetal bovine serum (Gibco) in Eagle’s Minimum Essential Medium (MEM, Corning) with 1% penicillin/ streptomycin (Thermo Fisher Scientific). Cells were cultured at 37 °C with 5% CO2 and passaged at around 80% confluency. SH-SY5Y cells were differentiated into neurons as previously described with a few modifications as follows ^32^. A day after plating, the cells were infected with the virus. 48 hours post-infection, the cells were induced with fresh differentiation medium containing serum-free neurobasal/B27 plus culture medium (Thermo Fisher Scientific), Glutamax (Thermo Fisher Scientific), penicillin/ streptomycin, 50 ng/ml BDNF (Alomone Labs), 20 mM KCl, 2 mM dibutyryl cyclic AMP (db-cAMP), and 10 μM all-trans retinoic acid for 18 days.

### Total RNA extraction and RT-qPCR

For fetal lung tissues and peripheral blood mononuclear cell (PBMC), total RNA was extracted with QIAGEN miRNeasy® Mini kit according to the manufacturer’s instructions. For cell lines, total RNA was extracted with TRIzol (Invitrogen) and treated with DNase I (NEB) according to the manufacturer’s instructions. The RNA was reverse transcribed with 50 ng random hexamers using Superscript III Reverse Transcriptase (Invitrogen) in a 20 μL reaction for 10 min at 25 °C followed by 50 min at 50 °C. The complementary DNA (cDNA) was PCR amplified using forward and reverse primers containing sequences specific to the snoRNAs (listed in **Table S2**). In a 20 μL reaction, 3 ng of cDNA template, 0.5 μM each of forward and reverse primers, 10 μL Power-up Universal SYBR Green 2× Supermix (Applied biosystem) were mixed with nuclease-free water and amplified for 40 rounds at an annealing temperature of 60°C on a StepOnePlus Real-Time PCR System (Applied Biosystems). Mean cycle threshold (CT) values were determined for control and experimental samples. Changes in snoRNA expression were normalized to hMRP mRNA expression level using the 2−ΔΔCT method.

### RNA-seq library preparation and sequencing

The rRNA-depleted RNA-seq libraries were prepared according to previously described protocols.^33^ Briefly, the Ribo-Zero rRNA Removal Kit was used to deplete rRNAs, and the NEBNext UltraTM RNA Library Prep Kit was used to prepare libraries. qPCR was used to quantify library concentration; the Tapestation High Sensitivity D1000 Screen Tapes were used for library quality control. Libraries were then pooled and sequenced on the NovaSeq X Plus platform with 150 PE read lengths, and a target of 80M total reads (40M in each direction) per sample (Admera Health LLC).

### RNA-seq analysis

RNA-seq reads were first mapped to human genome hg38 with published General Transfer Format (gtf) files, including all types of RNAs using Tophat 2 version 2.1.0.^34,35^ RNA-seq reads from ZNHIT3 Δ251-254 were also mapped to the human genome hg38 using STAR to confirm the expression of exogenous ZNHIT3.^36^ Differential expression (DE) analysis was performed by cufflinks version 2.2.1, and genes with adjusted *p* value less than 0.05 were considered significant DE genes.^37^ Gene Ontology analysis was performed using PANTHER release 8.0 and visualized by ggplot2.^38^

### Analysis of global 2′-O-methylation levels by reverse transcription

The level of 2′-O-methylation of 28S rRNA was assayed in three large regions as previously described.^39^ Briefly, 1 μg total cell RNA extracted using the TRIzol method was treated with DNase I (NEB), then reverse transcribed with 50 ng random hexamers in a high dNTP concentration (10 mM) or low dNTP concentration (0.1 mM) using 200 U SuperScript III reverse transcriptase (Invitrogen) in a 20 uL reaction for 50 min at 50 °C. Samples were treated with RNase H, and cDNAs were analyzed by qPCR using the PowerUp SYBR Master Mix (ThermoFisher) with the oligos listed in **Table S2**. The following thermocycler setup was used in a CFX Opus 96 instrument (Biorad): 2 min at 50°C followed by 40 cycles of 15 s at 95 °C, 30 s at 60 °C, and 30 s at 72 °C. The quantification cycle delay for each region was calculated as follows: ΔCq = low dNTP Cq – high dNTP Cq. This value was normalized for each region to the threshold cycle delay of the unmethylated 28S region (Region 1).

### RiboMethSeq analysis of rRNA modifications

The RiboMethSeq analysis, which measures 2’-O-methyl-induced protection of the RNA phosphodiester bond against alkaline hydrolysis, was performed as previously described, using the validated list of Nm positions present in *H. sapiens* rRNA.^40,41^ Briefly, 100 ng of total RNA isolated from lung tissue samples from the second affected fetus and two age-matched controls, as well as from parental peripheral blood mononuclear cell (PBMC) RNA, were subjected to alkaline hydrolysis at 96 °C for 12 min. The precipitated and end-repaired RNA was then converted into a library using the NEBNext© Small RNA Library Kit (NEB). The quality and quantity of the library was assessed using a High Sensitivity DNA Chip on a Bioanalyzer 2100 and a Qubit 2.0 fluorometer. High-throughput sequencing of the multiplexed libraries was performed on an Illumina NextSeq 2000 instrument in a 50 nt single end read mode.

### Analysis of global translation by puromycin incorporation

Translation capacity was measured by quantifying the newly synthesized peptides using a puromycin incorporation assay in HEK293T cells transfected with wild-type or ZNHIT3 variants. Puromycin was added to the media 48 hours post-transfection to a final concentration of 10 μM, and cells were incubated for 60 min at 37 °C. The cells were washed with ice-cold PBS and lysed in RIPA buffer. Lysed samples were clarified by centrifugation at 13000 rpm for 10 min. Clarified samples were analyzed by Western blotting. Anti-puromycin antibody was used to measure the amount of puromycin incorporation.

### Antibodies

All antibodies used in this study were commercially available and include HA tag monoclonal antibody (Invitrogen; 26183), GAPDH monoclonal antibody (Proteintech; 60004-1-Ig), ZNHIT3 Polyclonal Antibody (Invitrogen; PA5-43577), ZHNIT3 Polyclonal Antibody (HPA078211, Sigma-Aldrich), Cyclophilin A Polyclonal Antibody (AF3589, R&D Systems), PIH1D1 Polyclonal Antibody (Proteintech; 19427-1-AP), RPAP3 Polyclonal Antibody (Proteintech; 23741-1-AP), NOP58 Polyclonal Antibody (Proteintech; 14409-1-AP), NUFIP1 Polyclonal Antibody (Proteintech; 12515-1-AP), NHP2L1 Polyclonal Antibody (Proteintech; 15802-1-AP), and anti-puromycin antibody (Sigma-Aldrich, MABE343).

### Data availability

RNA-seq data are deposited to NCBI’s Gene Expression Omnibus (GEO) and are available under accession code GSE273112. RiboMethSeq data are deposited to the European Nucleotide Archive (https://www.ebi.ac.uk/ena/browser/home) and are available under accession code PRJEB78345.

## Results

### Clinical findings

Healthy, unrelated parents in their late 20s were referred to the genetic consultation for multiple fetal losses. During the first pregnancy, fetal demise due to fetal hydrops occurred at 15-20 weeks of gestation (GW). Fetal pathology revealed a female fetus with normal growth parameters and no malformations but showed signs of placental hydrops. Neuropathology examination was limited. Chromosomal anomaly investigations were negative. In the second pregnancy, an early miscarriage occurred at 7-12 GW for which no fetal pathology was performed. The third pregnancy experienced a recurrence of fetal hydrops. The first-trimester ultrasound was normal, with a nuchal translucency of 1.7 mm and a crown-rump length of 60.9 mm. However, a morphologic ultrasound performed at 15-20 GW showed generalized edema and pericardial effusion. The medical termination of pregnancy took place at 17-22 GW. An autopsy revealed a slightly edematous male fetus with predominant edema in the extremities and slight retrognathia but no visceral malformations. Neuropathological examination showed measurements and maturation consistent with gestational age, without microscopic anomalies. The male fetus from the third pregnancy had slightly edematous appearance of the face and limbs, with more pronounced edema at the extremities and tapered fingers. **Figures 1A-B** show the macroscopic view of the brain after fixation for this fetus compared to a normal control fetus of the same age shown in **Figures 1C-D**. Microarray analysis and investigations for lysosomal storage diseases in the amniotic fluid sample were normal.

**Figure 1.**
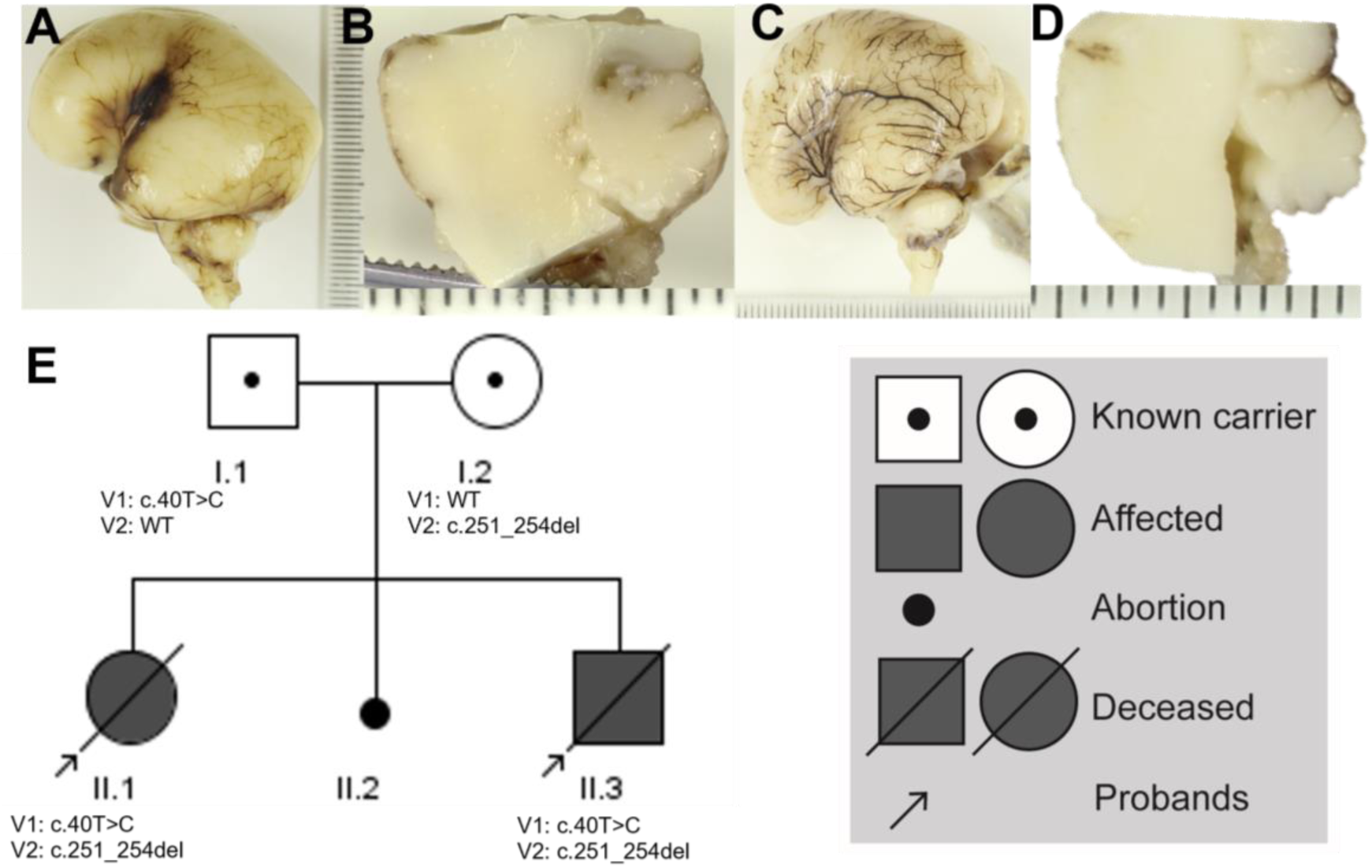
Macroscopic description of the two affected fetuses. (**A-D**) Macroscopic view of the fetal brains after fixation: (**A**) Left lateral view of the affected male fetus brain, (**B**) the midsagittal section of the cerebellum of the affected male fetus. **(C-D)** Similar views of a normal control fetus of the same age. **(E)** pedigree of the family with two affected offsprings. Shaded symbols indicate affected individuals. The genotype is shown for those individuals who were analyzed. All genotypic carrier individuals are phenotypically healthy.

### Molecular analysis

Whole-genome sequencing was performed on the fetuses from the first and third pregnancies and the parents. Two variants in the *ZNHIT3* gene were identified in both fetuses, each inherited from one parent (**Figure 1I**). The paternal variant NM_004773.4: c.40T>C, p.(Cys14Arg), located in exon 1, is reported at extremely low frequency in population control databases (gnomAD v4). All in silico prediction tools tested (REVEL, Polyphen 2, Alphamissense) supported its deleterious nature (CADD score:27.9). Moreover, another substitution affecting the same amino acid has been reported previously in a patient with PEHO syndrome.^29^ The maternal variant NM_004773.4: c.251_254delAAGA, located in exon 4, is a frameshift deletion of 4 nucleotides predicted to lead to a premature stop codon (p.(Glu84Alafs*8)) reported with a frequency compatible with recessive transmission in population control databases (gnomAD v4) (**Figure 3A-B**).

### Analysis of the expression levels of the newly identified ZNHIT3 variants in brain samples

To evaluate the effect of the two variants in combination on the localization and expression of ZNHIT3 in the fetal brain, we compared the expression of ZNHIT3 in brain tissue sections from one of the affected fetuses with that of two age-matched fetal controls (17-22 GW) by Immunohistochemistry on paraffin sections. The analysis was performed on the coronal brain (**Figure 2A-C**) and sagittal cerebellum (**Figure 2D-E**) sections. Haematoxylin and Eosin Staining (HES) showed no migration anomalies and normal foliation of the cerebellum, with different cell layers having similar appearances (**Figure S1**). Overall, the expression pattern of ZNHIT3 was similar in both affected individuals and controls, with strong cytoplasmic staining observed in cells that appeared to be neurons. ZNHIT3 expression was detected in specific regions, including the ventral part of the corpus callosum, the fornix, the external and internal capsules, the optic tracts, and areas around the third ventricle near the thalamus. Clusters of strongly stained cells were found in the lateral and temporal horns of the cerebral ventricles within germinative zones. Additionally, in the brainstem, the germinative zone around the fourth ventricle also exhibited ZNHIT3 expression, similar to the expression seen in the cerebellar peduncles (**Figure 2F**).

**Figure 2.**
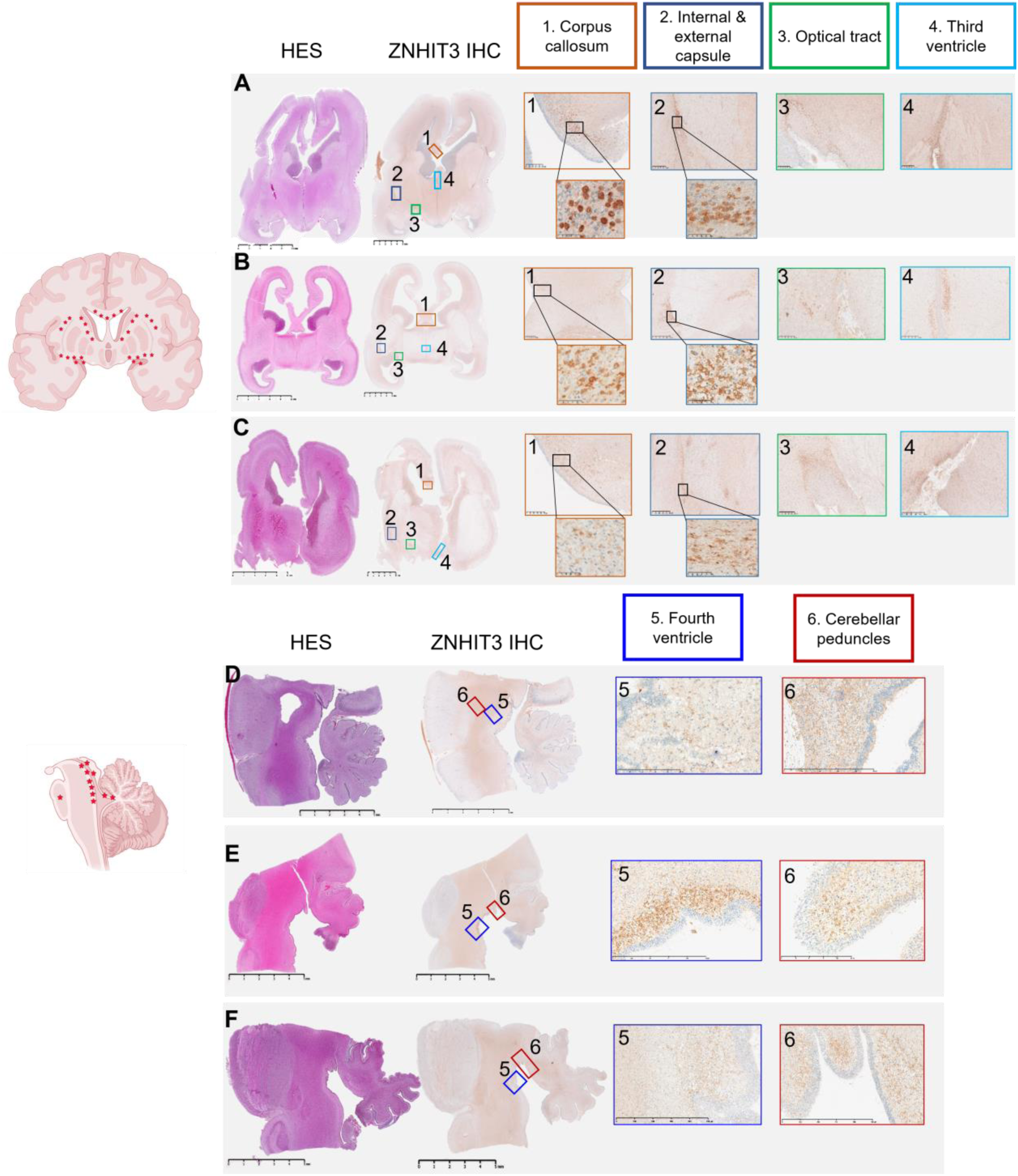
Analysis of ZNHIT3 expression in brain sections. ZNHIT3 expression was measured in the developing brain and cerebellum by immunohistochemistry on paraffin sections. **(A-C)** HES and IHC magnification of the coronal brain sections. **(D-F**) Sagittal cerebellum sections. **A** and **D** show the results for the affected male fetus, and **B** and **E** show the results for the age-matched control 1 and **C** and **F** for age-matched control 2. HES showed no migration anomalies and normal foliation of the cerebellum, with different cell layers having similar appearances. Purkinje cells are not yet identifiable at this stage of development. HES: Haematoxylin and eosin-stained, IHC: immunohistochemistry.

### Newly identified *ZNHIT3* variants affect protein stability and cell growth in cell line models

PEHO-associated *ZNHIT3* variants have been shown previously to affect the steady-state levels of the protein when modeled in yeast or human cell lines.^28,30^ To assess how the newly identified *ZNHIT3* variants affect the steady-state levels of the protein in growing cells, we introduced the HA-tagged wildtype or variant *ZNHIT3* into HEK293T cells. We then treated cells with cycloheximide and assessed the protein level after 0, 2, 4 and 6 hours by immunoblots using anti-HA antibody for detection of ZNHIT3 and anti-GAPDH as control. An empty vector transfection was also included as a control. Immunoblots showed that the C14R variant reduces the protein levels significantly, whereas the Δ251-254 variant is more stable than the wild type (**Figure 3C**). In comparison, both previously reported PEHO-associated *ZNHIT3* variants destabilize the protein (**Figure 3D**).

**Figure 3.**
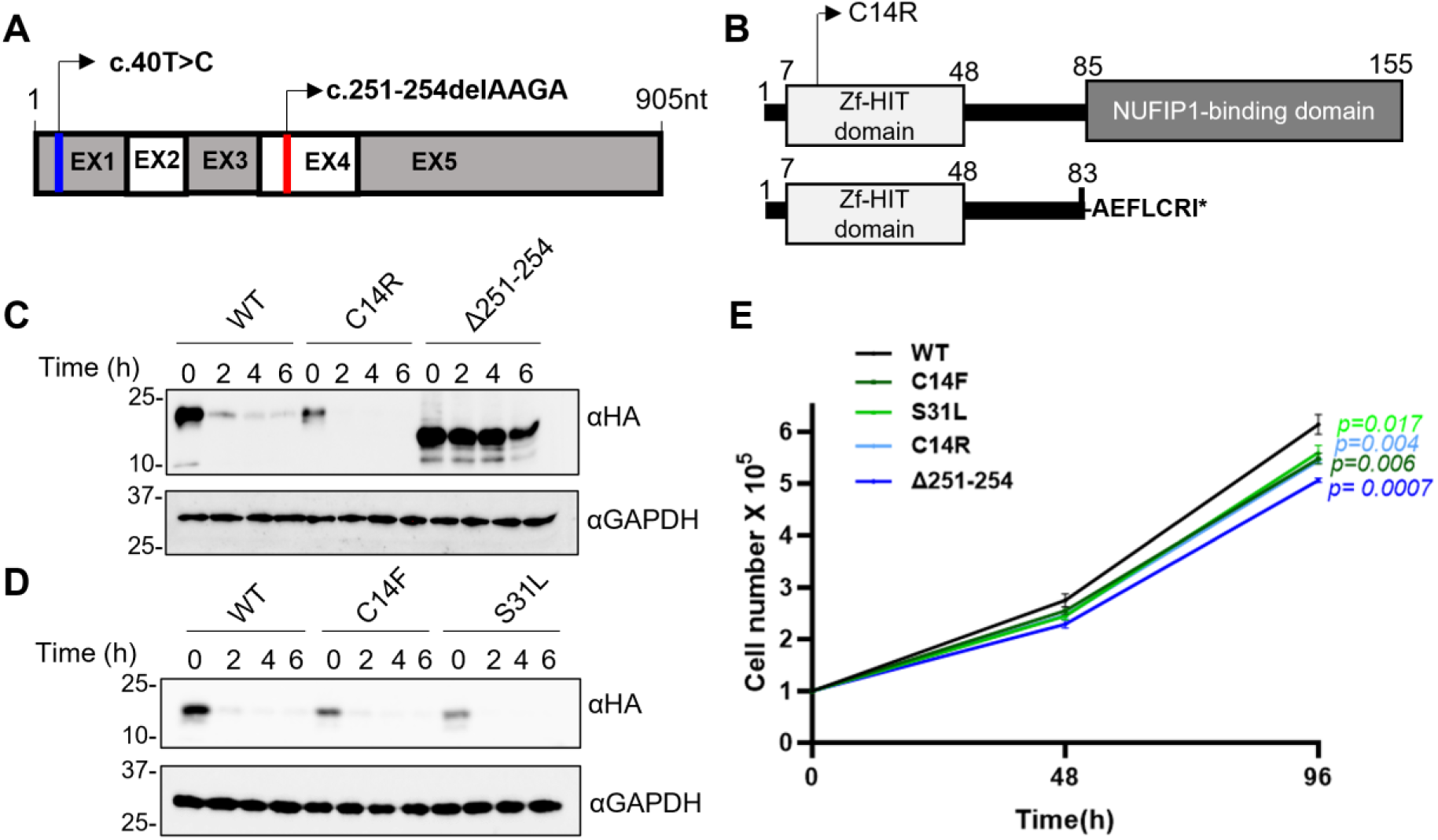
ZNHIT3 variants affect protein stability and cell growth. (**A**) Schematic of the location of the newly identified pathogenic ZNHIT3 variants. (**B**) Domain organization of ZNHIT3 protein and the new variants. **(C-D**) Immunoblots assaying the steady-state levels of ZNHIT3 and its variants. Cells were treated with cycloheximide (10 μg/ml) for 0, 2, 4, or 6 h, and an equal concentration of cleared total cell lysate was analyzed by immunoblotting. **E**) Assay of cell growth at days 0, 2 and 4 in cells expressing ZNHIT3 or its variants. Three independent biological replicates were transfected and measured. *p* values were calculated using an unpaired *t* test on data from the 96 h timepoints.

Deficiency of ZNHIT3 is shown to result in a reduced number of cultured mice cerebellar granule neurons.^28^ To investigate whether and how the newly identified *ZNHIT3* variants affect cell growth, we assayed the growth of HEK 293T cells expressing wild type and variants of *ZNHIT3*. Cells expressing *ZNHIT3* variants showed a significant decrease in their growth compared to wild-type control cells expressing an empty vector (**Figure 3E**). The impact from newly identified ZNHIT3 variants was comparable to the effect from S31L and C14F *ZNHIT3* variants described previously,^29^ while the Δ251-254 variant had a greater detrimental effect on cell growth than other variants. Thus, the newly identified ZNHIT3 variants have different effects on the steady-state levels of the protein by either destabilizing (C14R) or stabilizing (Δ251-254) the protein, but both impact cellular growth.

### Expression of *ZNHIT3* variants causes substantial gene expression changes

To check if cellular gene expression is affected by the newly identified *ZNHIT3* variants, we performed an unbiased total RNA-seq analysis. For this assay, we took advantage of SH-SY5Y cells, a well-characterized neuroblastoma cell line for which differentiation protocols are established.^32^ Undifferentiated SH-SY5Y cells were transfected with lentiviral plasmids expressing wild type or variant *ZNHIT3*. Total RNA from undifferentiated cells at day 0 and differentiated cells at day 18 were extracted and analyzed by RNA-seq. Over 90% of total *ZNHIT3* transcripts contained the expressed variant and the undifferentiated and differentiated cells were clearly distinguishable by their RNA expression profiles. **Figure 4A** depicts differences in gene expression between cells expressing each new *ZNHIT3* variant compared to wildtype control in differentiated and undifferentiated cells. Expression of *ZNHIT3* variants caused distinct and shared gene expression changes (**Figure 4B**). Transcriptomic changes were more significant in cells expressing *ZNHIT3* Δ251-254 variant than those expressing *ZNHIT3* C14R variant both on day 0 and day 18. Despite higher overall protein stability of ZNHIT3 Δ251-254 variant, this protein lacks a critical domain required for its interaction with NUFIP1.

**Figure 4.**
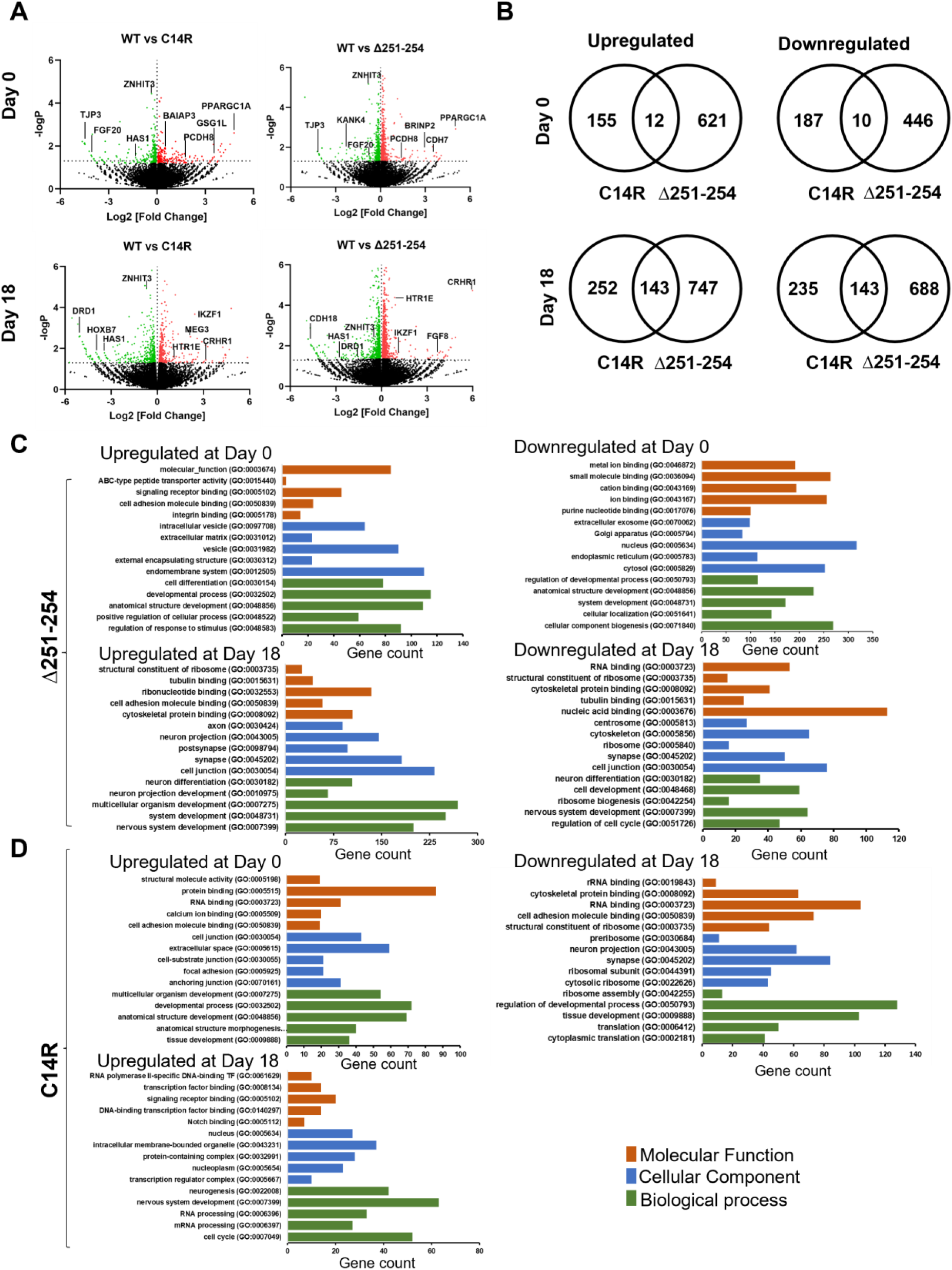
ZNHIT3 variants cause gene expression changes. **(A)** Differential gene expression analyzed by RNA-seq. Total RNA was isolated from SH-SY5Y cells expressing wildtype or variant ZNHIT3 at undifferentiated (day 0) and differentiated (day 18) states and was depleted for rRNA before library prep and sequencing. Volcano plots of differential gene expression data were generated in GraphPad Prism using log_2_ fold change and -log_10_ adjusted p-value. Non-significant changes (adj p-value> 0.05) are indicated in *black* and fall under the horizontal dashed line. Datapoints on the left side of the *x*-axis indicate significantly downregulated genes and those on the right indicate significantly upregulated genes. (**B)** the statistically significant upregulated and downregulated genes in cells expressing C14R or Δ251-254 identified by RNA-seq are compared using a Venn diagram. (**C-D**) Gene ontology (GO) analysis of the significantly upregulated and downregulated transcripts was performed using Panther and categorized by biological process, cellular component and molecular function.

Only twelve upregulated and ten downregulated genes were commonly shared between the two variants in the undifferentiated state at day 0, highlighting the distinct effects of these variants. Many of the differentially expressed genes in undifferentiated cells were linked to cell signaling pathways, including *FGF20*, *TJP3*, and *PPARGC1A*, depicted in **Figure 4A**.^42-44^ In the differentiated cells, 143 genes were shared between the two variants, including genes involved in ribosome biogenesis, such as *HAS1*, and others involved in neurodevelopment, such as *DRD1*, *CRHR1*, and *HTR1E*, depicted in **Figure 4A**.^45-48^ GO term analysis of upregulated transcripts in undifferentiated cells expressing the *ZNHIT3* Δ251-254 variant revealed that most differentially expressed transcripts are related to regulating developmental processes, whereas downregulated genes are linked to cellular component biogenesis. Similarly, upregulated genes in undifferentiated cells expressing the *ZNHIT3* C14R are associated with developmental-related pathways, however, downregulated genes did not fall into significantly enriched GO terms. In differentiated cells expressing either of the *ZNHIT3* variants, downregulated transcripts are linked to ribosome biogenesis and translation, whereas upregulated genes are mostly involved in nervous system development, neurogenesis, and differentiation pathways (**Figure 4C-D**). In summary, the RNA-seq data reveal distinct gene expression changes in cells expressing the new *ZNHIT3* variants and show downregulation of ribosome biogenesis and translation as a commonly affected pathway during differentiation in these cells.

### The newly identified *ZNHIT3* variants affect the steady-state levels of snoRNP assembly factors and snoRNP biogenesis

To evaluate the impact of *ZNHIT3* variants on ribosome biogenesis and translation, we focused on the known function of ZNHIT3 in snoRNP assembly. We assessed if the new ZNHIT3 variants impact snoRNP assembly by altering the steady-state levels of assembly factors and/or the ability of the variants to form complexes. To test how *ZNHIT3* variants impact the steady-state levels of other snoRNP assembly factors, we transfected HEK293T cells with HA-tagged wild-type or variant ZNHIT3, collected semi-confluent cells and assayed the level of each protein by immunoblotting. This analysis showed that a decrease in the level of ZNHIT3 by the C14R variant reduces the steady-state levels of NUFIP1 protein (**Figure 5A-B**). In contrast, expression of the stable ZNHIT3 Δ251-254 variant increases the steady-state level of NUFIP1. PIH1D1 steady-state levels followed a similar trend with a statistically significant higher steady-state level of PIH1D1 in cells expressing the Δ251-254 variant (**Figure 5A-B**). In contrast, the steady-state levels of RPAP3, 15.5K, and Nop58 did not change significantly. These data demonstrate that the C14R and Δ251-254 variants likely exert their impacts through different mechanisms, influenced by their protein different stabilities.

**Figure 5.**
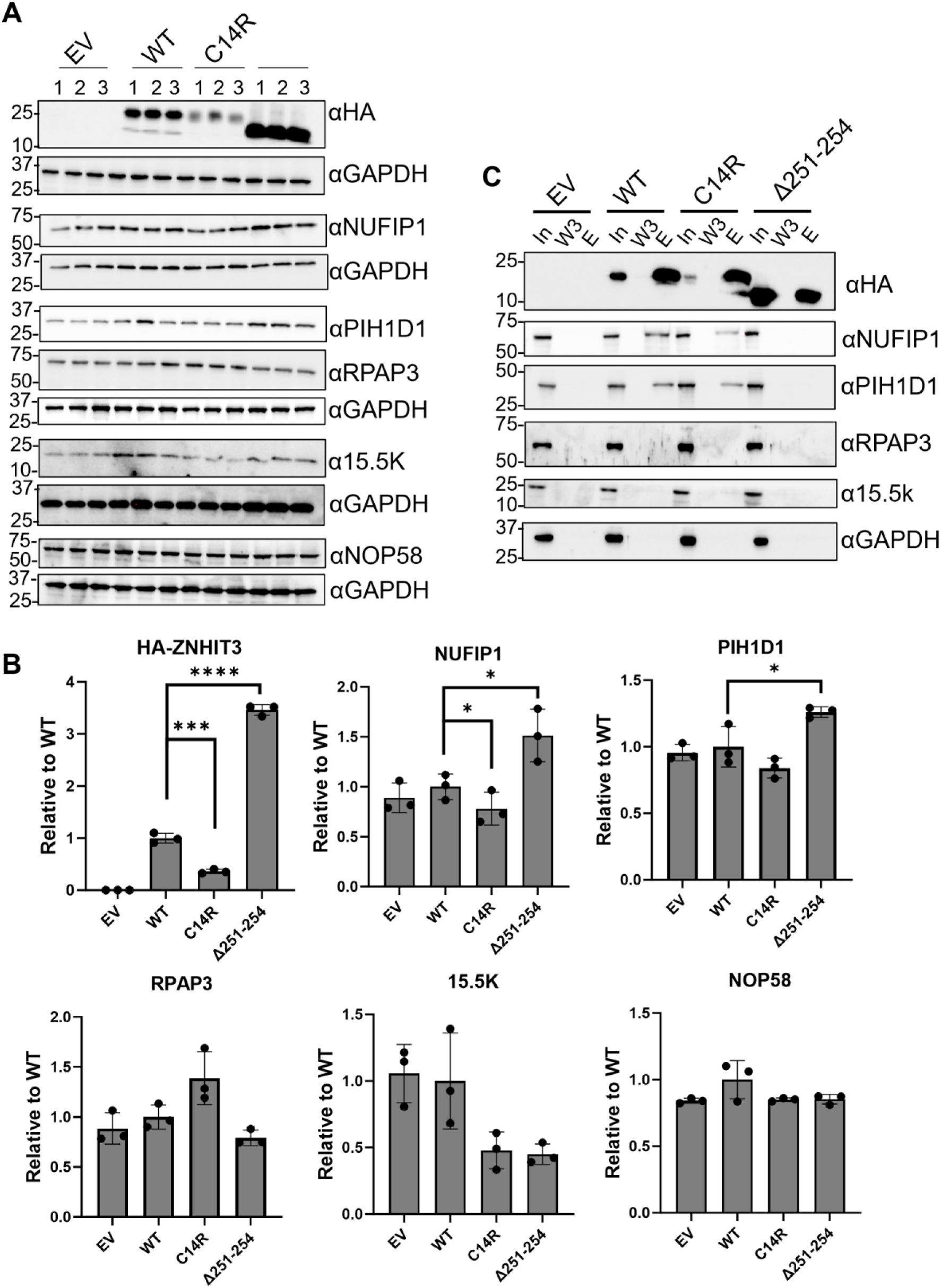
Expression of ZNHIT3 variants alters the steady-state levels of snoRNP assembly factors and impacts snoRNP assembly. **(A)** Immunoblots assaying the steady-state levels of proteins involved in snoRNP biogenesis in cleared total cell lysates of HEK293T cells expressing an empty vector (EV), or N-terminally HA-tagged ZNHIT3 (WT) or ZNHIT3 variants (C14R or Δ251-254). (**B)** Quantification of immunoblots shown in A. Data are normalized to GAPDH signal from each immunoblot transfer and are plotted relative to WT. Significance was determined using an unpaired *t* test comparing WT and each variant. For all graphs, bars represent the mean and standard deviation of three independent biological replicates. ∗*p* < 0.05. **(C)** Co-immunoprecipitation of HA-ZNHIT3 and variants (C14R or Δ251-254). Immunoblot analysis of core and assembly factors involved in snoRNP biogenesis. Analysis shows the effect on the interaction of ZNHIT3 with NUFIP1 & PIH1D1 proteins. In: input, W3: last wash, E: Eluate.

To test the effect of ZNHIT3 variants on the protein’s interaction with snoRNP assembly factors, we performed immunoprecipitation to isolate complexes using HA-tagged wild-type or ZNHIT3 variants in HEK293T cells. As control, we transfected cells with an empty vector expressing the HA-tag, but no ZNHIT3. In cells, expressing wild-type HA-tagged ZNHIT3, we isolated complexes containing NUFIP1 and PIH1D1. As anticipated, the ZNHIT3 Δ251-254 variant that lacks the NUFIP1-binding domain did not interact with NUFIP1 (**Figure 5C**). Compared to wild-type ZNHIT3, less NUFIP1 and PIH1D1 were pulled down by the ZNHIT3-C14R variant which destabilizes the protein. We did not detect ZNHIT3 interactions with other tested snoRNP assembly factors or core proteins under these conditions with either wildtype of variant ZNHIT3 (**Figure S2**). Collectively, these data indicate that the ZNHIT3 Δ251-254 variant is unable to interact with NUFIP1 and PIH1D1 thereby limiting the ability of the protein to contribute to snoRNP assembly. Whereas the ZNHIT3-C14R variant can participate in snoRNP assembly, albeit less efficiently.

### The new *ZNHIT3* variants cause site-specific hypo rRNA 2’-O-methylations

Impaired C/D snoRNP assembly in yeast models of PEHO-associated ZNHIT3 variants results in site-specific rRNA 2’-O-methylation changes.^39^ To assess whether the expression of new ZNHIT3 variants may have a similar effect in human cells, we first employed a reverse transcription-based assay to globally analyze the 2′-O-methylation of the 28S rRNA in total RNA that was isolated from ZNHIT3 variants transfected HEK293T cells. We performed this analysis on three regions of the 28S rRNA with 8, 14, or 12 2′-O-methylations, respectively, and used a region without any rRNA 2′-O-methylations as an internal control for normalization (**Figure 6A**). The result from this analysis shows a significant hypo 2′-O-methylation in all three tested regions of rRNA isolated from cells expressing the Δ251-254 mutant (**Figure 6B**).

**Figure 6.**
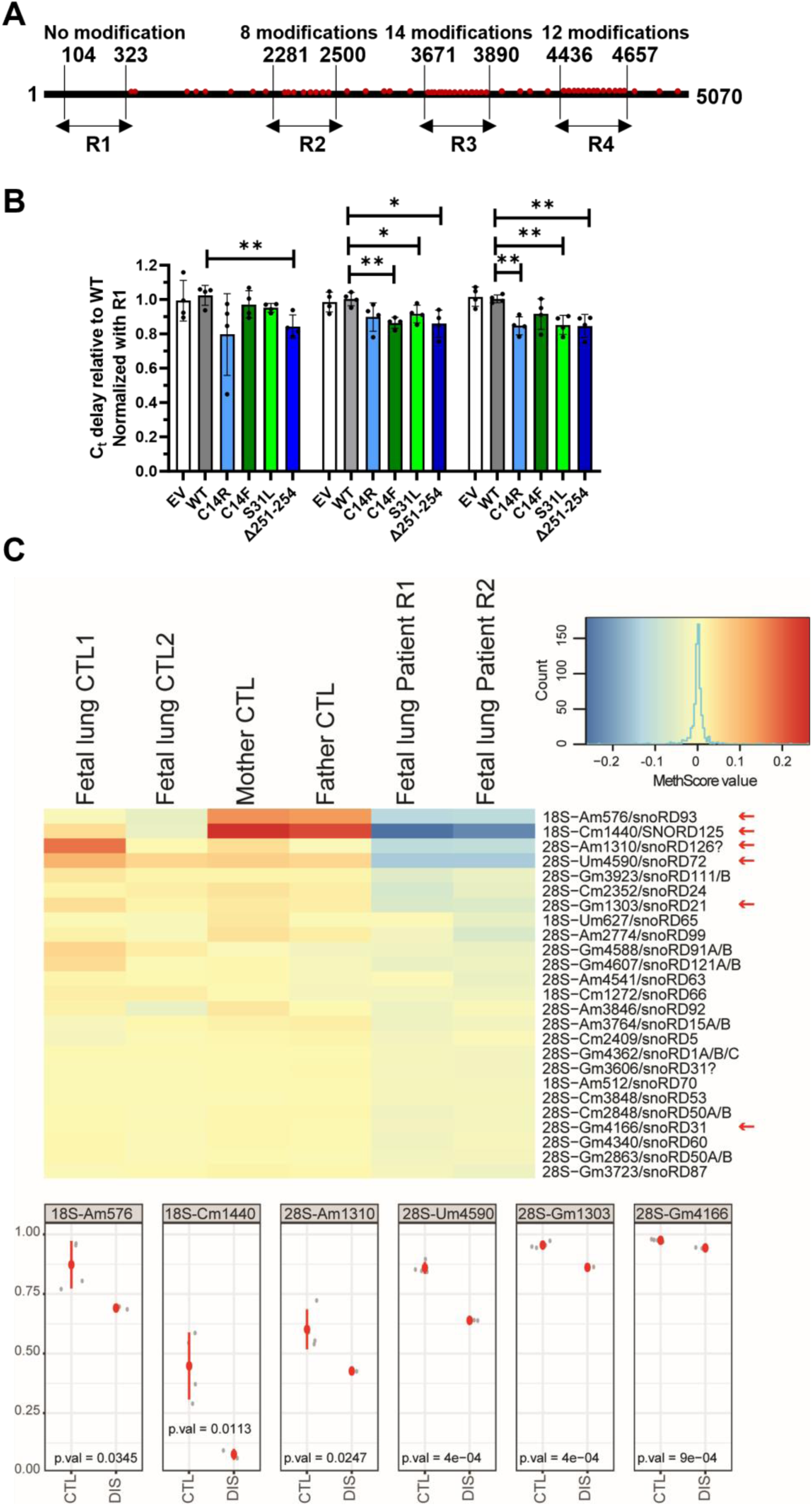
The new ZNHIT3 variants cause site-specific hypo rRNA 2’-O-methylation. **(A)** schematic of regions of 28S rRNA probed in B. **(B)** total RNA from HEK293T transfected cells was extracted, and three regions of 28S rRNA (regions 2–4) were probed for their 2′-O-methylation level by reverse transcription at low dNTP concentration followed by qPCR. Total RNA isolated from three independent biological replicates was used for the assay. Region 1, with no 2’-O-methylations, was used for normalization. Bars represent the mean and standard deviation of at least three independent biological replicates. P values were calculated by unpaired, Student’s t test. ∗*p* < 0.05; \*\**p*<0.01. **(C)** Partial differential heat-map of RiboMethSeq. MethScores for the most affected rRNA positions. MethScores are normalized to the mean by position with the color code in indicated to the right. Identity of rRNA Nm residue and associated box CD snoRNA (if known) is indicated. Analysis of rRNA 2’-O-methylation is shown for two control fetuses, from parents and from two replicates of affected fetal sample. Variation of the MethScore (Methylation level) for positions shown by arrows is illustrated at the bottom, CTL samples include both parental samples and irrelevant control fetuses, DIS condition is composed by two affected replicates. Identity of the rRNA site is shown on the top of each panel. Red dot shows mean value, red bar the s.d. value, grey dots show values for individual samples. P-value was calculated with unpaired Student’s t-test.

To assess whether expression of ZNHIT3 variants in patient cells may have a similar effect, we performed RiboMethSeq analysis on total RNAs isolated from preserved lung tissue samples of the affected male fetus from the third pregnancy and two age-matched controls, as well as parental peripheral blood mononuclear cells (PBMC). This analysis revealed several site-specific hypo rRNA 2’-O-methylation sites in the patient sample compared to the parental and healthy fetuses samples (**Figure 6C**). The most significantly hypomodified sites are 18S-Am576 and 18S-Cm1440 in the small ribosomal subunit and 28S-Gm1303, 28S-Am1310, 28S-4166, and 28S-Um4590 in the large ribosomal subunit. The complete profile of rRNA 2’-O-methylation pattern of the patient and controls is shown in **Figure S3A-B**.

### The new *ZNHIT3* variants reduce steady-state levels of specific snoRNAs

Deletion of the yeast *HIT1* gene or depletion of the human *ZNHIT3* gene both cause a reduction in steady-state levels of snoRNAs.^22^ Similarly, studies in yeast have shown that PEHO-associated *ZNHIT3* mutations reduce steady-state levels of box C/D snoRNAs.^30^ We, therefore, examined the extent to which the new variants of *ZNHIT3* in human model cells affect box C/D snoRNA levels. Using RT-qPCR, we quantified the levels of a panel of snoRNAs in cells expressing *ZNHIT3* variants compared to wild-type control cells (**Figure S4**). The panel P included C/D snoRNAs guiding modifications that were altered in patient cells as measured by RiboMethSeq, C/D snoRNAs guiding modifications that were not changed, as well as orphan C/D snoRNAs and C/D snoRNAs involved in rRNA processing. As control, we tested the level of candidate H/ACA snoRNAs and 7SL RNA, which are not expected to be regulated by ZNHIT3. For normalization, we used the hMRP RNA, which is not known to be dependent on snoRNP assembly. Our data show that the steady-state levels of some methylating C/D snoRNAs, but not all, were significantly reduced in cells expressing *ZNHIT3* variants (**Figure S4A**).

Among snoRNAs that guide 2’-O-methylations that were altered in patient samples (shown by arrows in **Figure 6C**), the steady-state levels of SNORD72, 93, 125 and 126 were lower in cells expressing the variants, in line with significant changes in their guided rRNA methylation status in fetus samples. The reduction in snoRNA levels appears to be specific to some methylating box C/D snoRNAs. Tested box H/ACA snoRNAs, C/D snoRNAs involved in rRNA processing that guide, orphan C/D snoRNAs, and 7SL RNA levels were not significantly changed relative to wild type (**Figure S4B**). To assess why certain snoRNA levels are more impacted than others, we compared snoRNAs for the length of their guide region, the energy of folding (ΔG (kcal/mol)), and their GC content and apparent melting temperature (Tm) of the guide region for methylating C/D snoRNAs (**Table S3**).^49^ This analysis shows that guides within SNORDs with altered steady-state levels have lower GC content and Tm values, potentially explaining the observed changes in snoRNA levels.

### *ZNHIT3* variants decrease cellular translation

Because *ZNHIT3* variants caused a change in gene expression related to ribosome biogenesis and translation (**Figure 4**) and impacted the steady-state levels of select snoRNAs and rRNA 2’-O-methylations (**Figures 6** and **S4**), we assayed whether global cellular translation is impacted by the new ZNHIT3 variants. To check how translational output is affected by ZNHIT3 variants in the human cell line models, we performed a puromycin incorporation assay and quantified global translation in bulk after 60 minutes. The incorporation of puromycin into newly synthesized proteins was quantified by western blots using an anti-puromycin antibody. The results showed that overexpression of all pathogenic ZNHIT3 variants in HEK293T cells decreases the translational output significantly relative to wild-type control cells (**Figure 7A-B, S5A**). RT-qPCR to measure the level of mature ribosomal RNAs in cells expressing ZNHIT3 variants compared to the wild-type control reveals a modest significant reduction in the steady-state level of 28S rRNA in cells expressing any pathogenic ZNHIT3 variants and a reduction in 18S rRNA in cells expressing C14R and Δ251-254 variants (**Figure S5B**). The steady-state levels of 5S and 5.8S rRNAs do not significantly change in cells expressing variants. Collectively, these data indicate that the expression of the newly identified ZNHIT3 variants reduces global translation and the steady-state cellular levels of mature 18S and 28S rRNAs.

**Figure 7.**
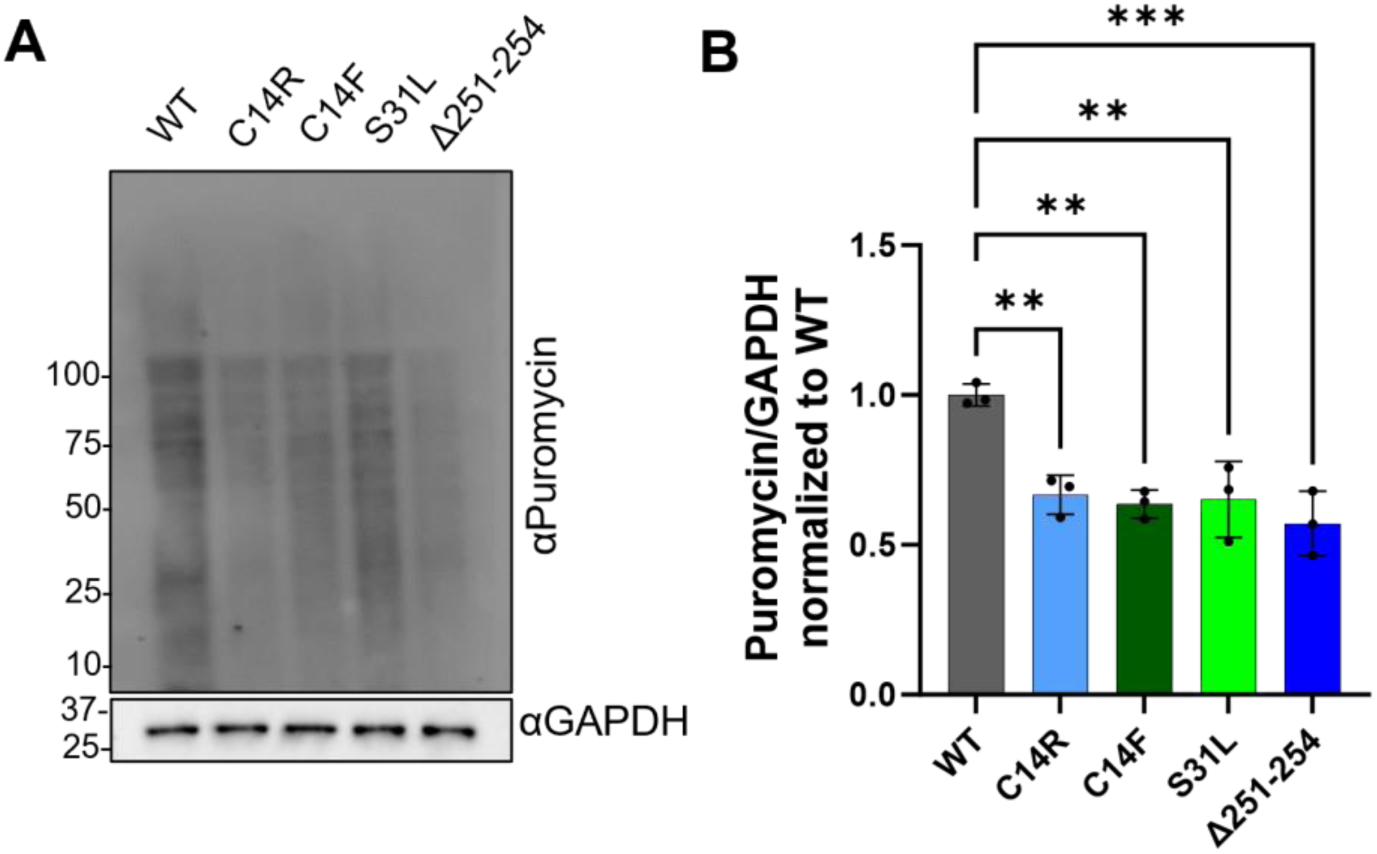
ZNHIT3 variants decrease cellular translation. **(A)** Puromycin incorporation assay was performed using an anti-Puromycin antibody in HEK293T cells after transfection with ZNHIT3 variants. A representative immunoblot is shown, indicating reduced protein synthesis in cells expressing ZNHIT3 variants. **(B)** Quantification of puromycylation immunoblots, using data shown in panel A and supplemental figure S5, from three independent biological replicates. Total lane intensity was normalized to the GAPDH signal per lane. Graph bars show mean values and error bars represent SD. *p* values were calculated using an unpaired, Student’s t test. \*\**p* < 0.01; \*\*\**p* < 0.001.

## Discussion

PEHO syndrome is an early childhood onset rare neurological disorder that is most common in the Finnish population in which a founder homozygous missense substitution p.(Ser31Leu) accounts for all the cases. To date, only one other *ZNHIT3* missense variant has been identified in a non-Finnish patient with PEHO syndrome. In this study, we report the first antenatal cases of PEHO syndrome in two non-Finnish siblings due to novel compound heterozygous variants in *ZNHIT3* and characterize their molecular defects. To our knowledge, this is the first reported instance of antenatal onset of PEHO syndrome resulting in early hydrops and intrauterine fetal loss. Our results suggest that this more severe phenotype may be associated with the Δ251-254 variant, which has more deleterious effects than previously reported missense variants in cell line models. Indeed, we show striking differences in the impact of the two variants C14R and Δ251-254 on the steady-state levels of the ZNHIT3 protein. The Δ251-254 variant produces a stable truncated ZNHIT3 protein with a loss of function in snoRNP biogenesis. In contrast, the C14R variant destabilizes the ZNHIT3 protein, reducing its cellular level while still supporting its function in snoRNP biogenesis, albeit at a reduced level. These findings support earlier studies on ZNHIT3 missense variants leading to PEHO syndrome, showing that the ZNHIT3-C14F and S31L also destabilize the protein. In addition, cells expressing the *ZNHIT3* Δ251-254 variant show a greater degree of cell growth defect and gene expression changes, suggesting the potential dominant negative effect of the truncated *ZNHIT3* protein variant.

Impairment of ZNHIT3 in zebrafish larvae induces microcephaly, structural cerebellar defects with granule cell defects, as well as pericardiac edema.^28^ Edema was present in the fetuses as early as 15 WG. However, no neurological abnormalities were observed on histological examination of the brain of the affected fetus. The expression pattern of ZNHIT3 observed in fetal brain tissues appears to be in accordance with the brain alterations seen postnatally in patients. Indeed, we observed strong expression in the germinative zones of the lateral ventricles and brainstem, as well as in the corpus callosum and cerebellar peduncles. These sites of expression are in line with the phenotype of progressive microcephaly with brainstem and cerebellar atrophy, associated with hypoplastic corpus callosum. Similarly, the optic tracts showed strong expression of ZNHIT3, which may contribute to the optic tract atrophy observed in PEHO syndrome.^27,28^ In mice, expression of ZNHIT3 is evident in proliferating fetal granule cell precursors of cerebellum at embryonic Day 16.5, and in proliferating and post-mitotic granule cells of cerebellum at postnatal Days 3 and 10.^50^ In murine development, the period from E15 to P10 is widely recognized as equivalent to the developmental stage of the third trimester of pregnancy in humans.^51^ Therefore, the specific neurological effects of ZNHIT3 are likely to occur late in neurodevelopment. Thus, the fetuses examined at 15 WG would be at a stage of development too early to see any neurological phenotype. Indeed, microcephaly and cerebellar atrophy are described as progressive and postnatally occurring features in PEHO syndrome.^27^

We analyzed gene expression changes in cells expressing ZNHIT3 variants compared to wildtype control because ZNHIT3 is also implicated in transcriptional regulation.^52-54^ In the analysis of differentiated and undifferentiated cells, the ZNHIT3 Δ251-254 variant showed a greater degree of differentially expressed genes than the ZNHIT3-C14R variant. This could be attributed to the loss of interaction of ZNHIT3 Δ251-254 with its binding partner NUFIP1, which is also implicated in transcription regulation.^55^ The observed differences in gene expression between differentiated and undifferentiated cells suggest that the loss of function of ZNHIT3 has a greater impact on mature cells, which aligns with the postnatal cerebral phenotype. ZNHIT3/Hit1p are shown to regulate the steady-state level of NUFIP1/Rsa1p,^56^ and genetically interact with PIH1D1.^57^ Here, we show that the new ZNHIT3 variants can alter the cellular steady-state levels of NUFIP1 and PIH1D1. Importantly, the expression levels of none of these genes are significantly altered in cells expressing ZNHIT3 variants, both in the differentiated or undifferentiated states, suggesting that failure to assemble snoRNP complexes, can alter the cellular levels of snoRNP biogenesis factors.

In the analysis of snoRNA steady-state levels, we found that the expression of pathogenic ZNHIT3 variants has a non-ubiquitous effect on the steady-state level of snoRNAs. Why the steady-state levels of some snoRNAs are more impacted by ZNHIT3 variants than others remain to be further investigated. A possible scenario is that snoRNAs with the shortest half-life due to their intrinsic properties and the nature of interactions with their target sites are mostly impacted by defects in snoRNP biogenesis. This scenario is supported by the observation that the steady-state levels of snoRNAs involved in rRNA processing that have extensive base pairing with rRNA are not impacted by ZNHIT3 variants. In contrast, our analysis of the short list of impacted snoRNAs suggests that the lower strength of snoRNA・rRNA base pairing can influence the stability of snoRNAs.

In summary, this study provides the first instance of antenatal onset of PEHO syndrome resulting in early hydrops and intrauterine fetal loss. Our work in cell line models defines vast molecular defects in gene expression, snoRNP biogenesis, and translation caused by the new *ZNHIT3* variants. A clear knowledge gap remains in understanding how the molecular defects caused by *ZNHIT3* variants result in specific developmental defects incompatible with life at a specific gestational age during pregnancy. Addressing this knowledge gap will also be relevant for other disorders caused by defects in ribosome biogenesis and translation that cause tissue-specific defects.

## Supporting information

Supplemtal Informaton

## Acknowledgements

The authors thank the patients’ family for their participation and cooperation in this study.

## Supplementary material

Supplementary material is included in a separate PDF file and is available online.

