## Supplementary material for "New *ZNHIT3* Variants Disrupting snoRNP Assembly Cause Prenatal PEHO Syndrome with Isolated Hydrops": Supplemtal Informaton

**Author affiliations:**

**Running title**: Fetal loss by novel *ZNHIT3* variants

**Keywords:** Intrauterine fetal loss; fetal hydrops; developmental disorder; PEHO syndrome; snoRNP biogenesis, translation

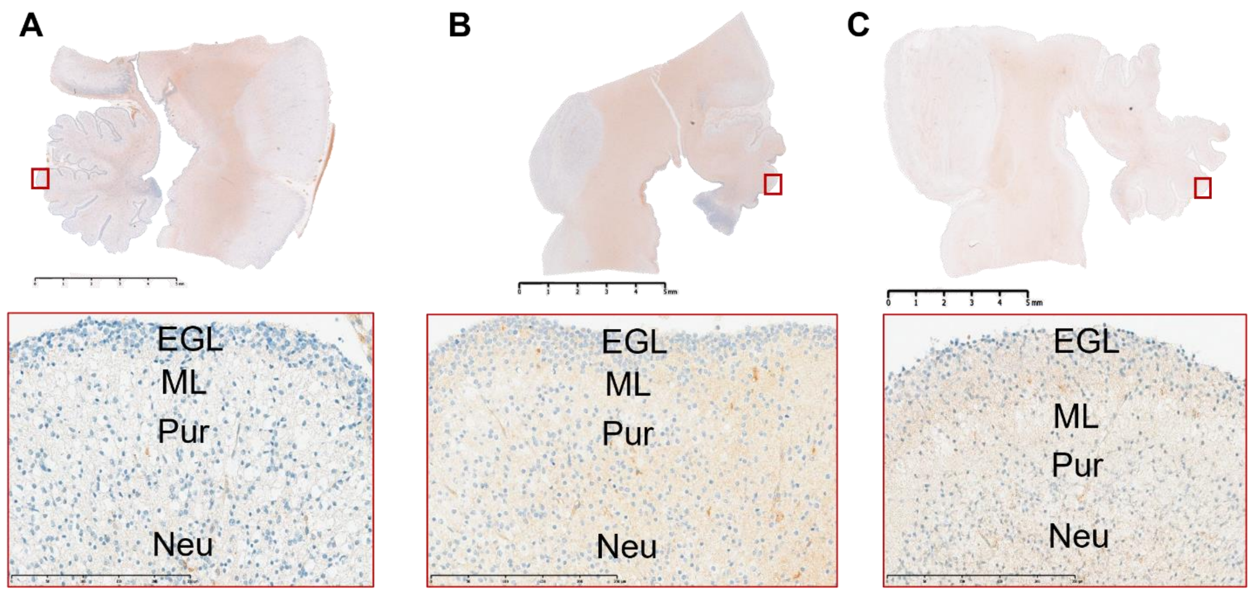


**Figure S1.** Immunohistochemical distribution of the ZNHIT3 protein in the cerebellum of the affected male fetus from the third pregnancy (**A**), control 1 at 15+4-GW (**B**), control 2 at 19+4 GW (**C**). External granular layer (EGL), thin molecular layer (ML), incomplete Purkinje layer (Pur), and migrated neuroblasts (Neu) are indicated. The internal granular layer was not detected. This figure is related to the main Figure 2.

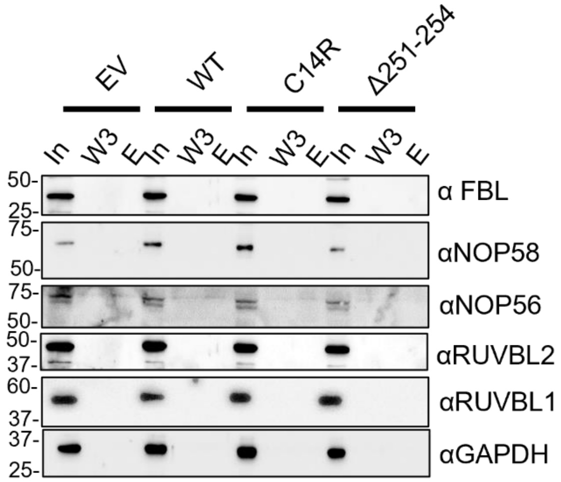


**Figure S2.** Immunoblots probing the interaction of new ZNHIT3 variants. Co-immunoprecipitation was performed using HEL293T cells transfected with HA-tagged wildtype or variant ZNHIT3, or an empty vector (EV) control just expressing the HA tag. Elution samples were purified via anti-HA tag magnetic beads. Data show no interaction of ZNHIT3 with core proteins FBL, NOP56 and NOP58 as well as assembly factors RUVBL1, RUVBL2 and a control protein, GAPDH. In=input, W3= wash 3 after binding, E= Eluate. This figure is related to the main Figure 5.

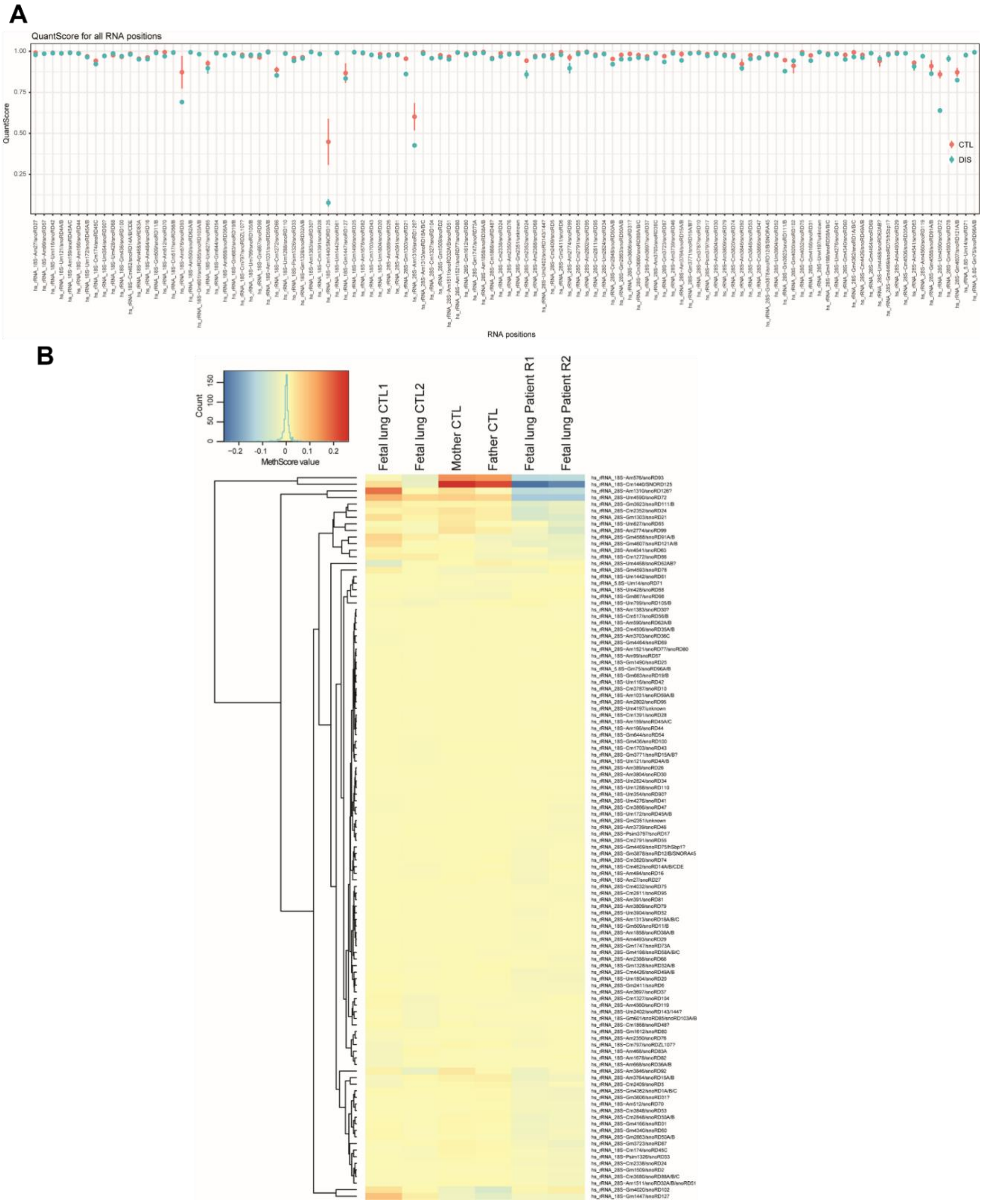


**Figure S3. (A)** Human rRNA methylation level (indicated as MethScore) for all rRNA modification sites. Mean and S.D. are shown in two groups (CTL) and disease (DIS). The identity of the site and the corresponding C/D snoRNA is shown at the bottom. **(B)** A full differential heat map showing MethScore variation for rRNA 2'-O-methylated positions in the fetus, parental samples, and fetal control samples is shown. This figure is related to the main Figure 6.

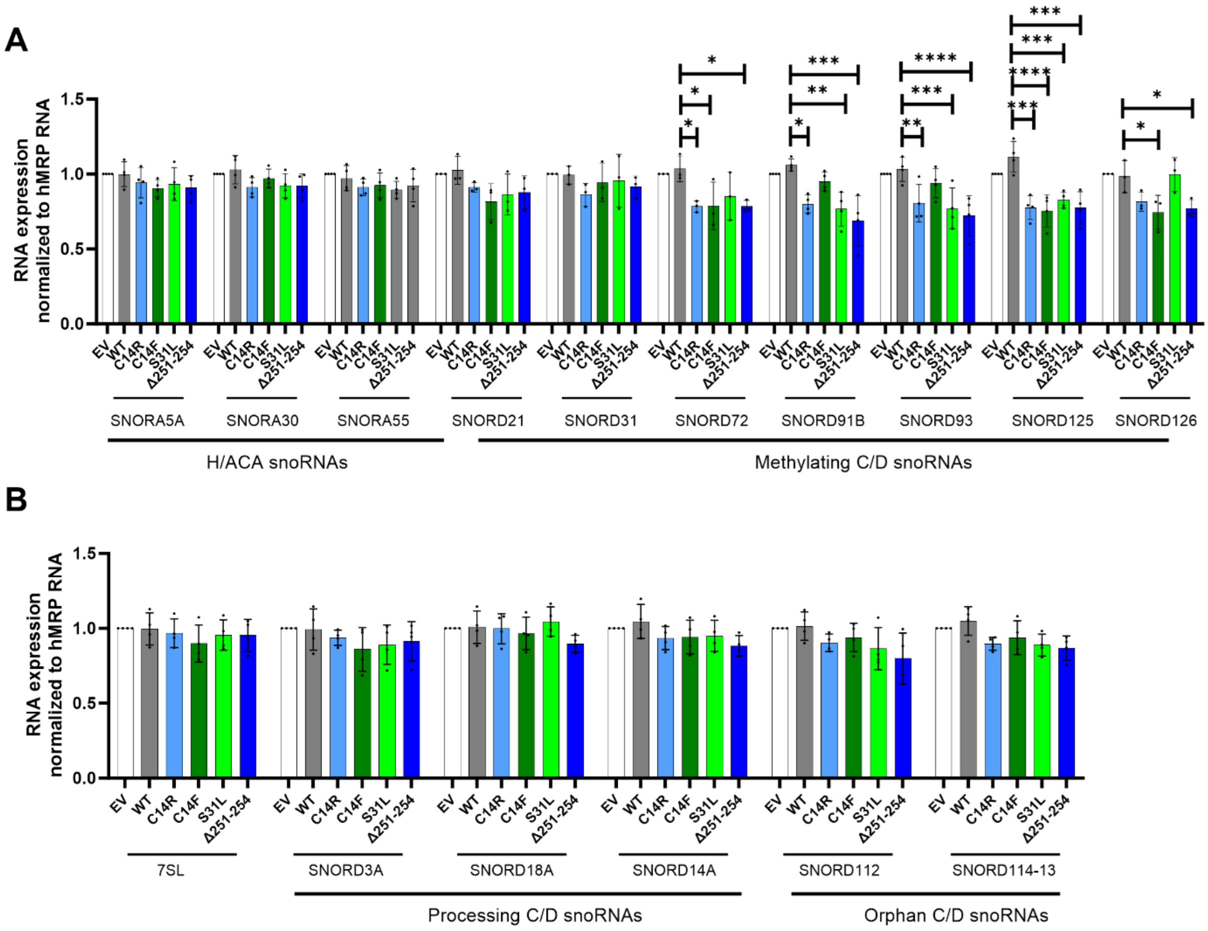


**Figure S4. The new ZNHIT3 variants decrease the steady-state levels of specific snoRNAs.** **(A)** Quantification of steady-state snoRNA levels of snoRNAs in HEK293T cells after transfection with an empty vector (EV), wildtype ZNHIT3 (WT) or ZNHIT3 variants (C14R, C14F, S31L, and Δ251-254) using RT-qPCR. The levels of seven box C/D snoRNAs were compared with three H/ACA snoRNAs. Data were normalized to hMRP RNA. **(B)** RT-qPCR quantification of steady-state snoRNA levels in HEK293T cells after transfection with ZNHIT3 variants. The levels of seven box C/D snoRNAs were compared with three H/ACA snoRNAs. Data were normalized to hMRP RNA. Bar graphs indicate the mean value and error bars show SD of six independent biological replicates. Significance was determined using an unpaired *t* test compared to wildtype control. $*p < 0.05$; $**p < 0.01$; $***p < 0.001$

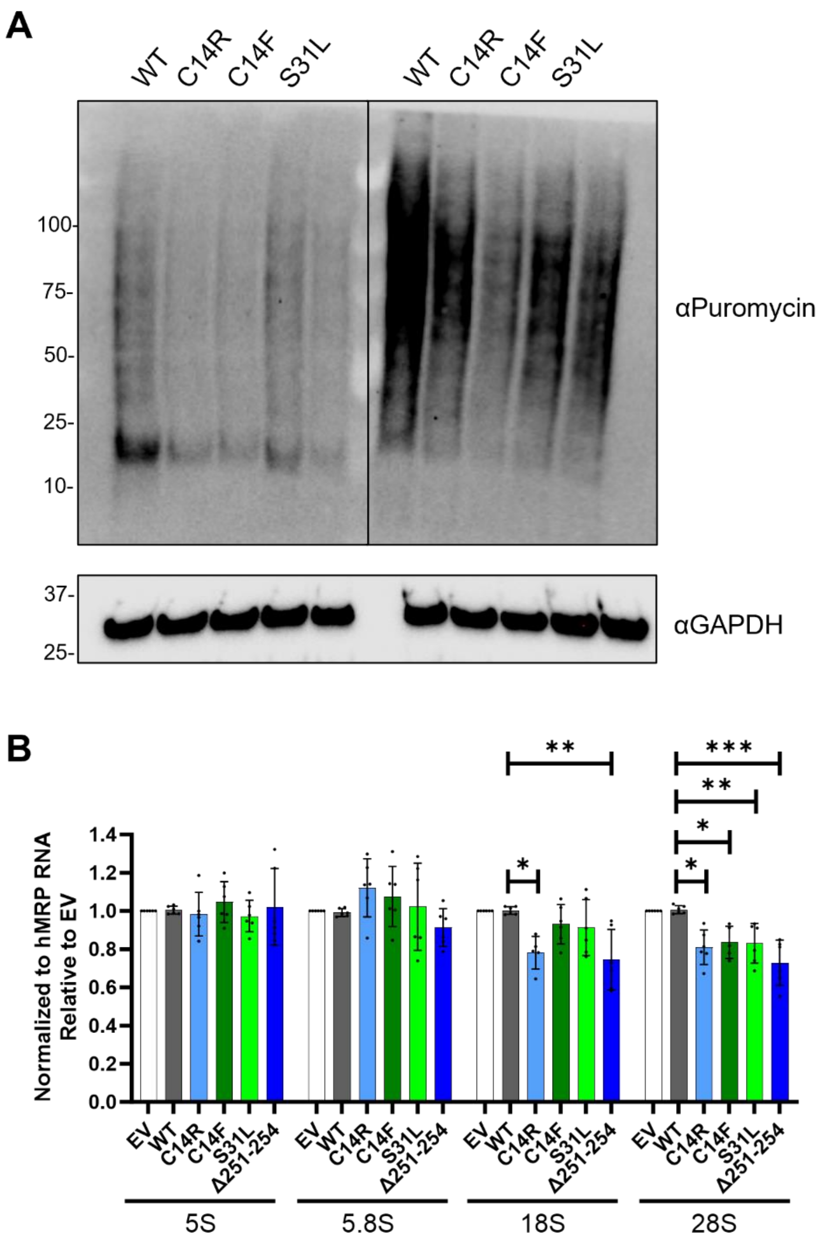


**Figure S5. Replicates of puromycylation assay.** **(A)** Puromycin incorporation assay was performed using an anti-Puromycin antibody. Two representative Western blots show reduced protein levels of cells expressing the ZNHIT3 variants. **(B)** RT-qPCR measuring steady-state rRNA levels. The experiment was carried out using RNA from 5-6 independent biological replicates. The bar shows the mean values and error bars represent SD. Significance was evaluated using an unpaired *t-test*. Asterisks indicate significant differences between WT and other variants. $*p < 0.05$; $**p < 0.01$; $***p < 0.001$. This figure is related to the main Figure 7.

**Table S1. List of plasmids used in this study.**

| Plasmid name | Backbone | Gene | Purpose |
|---|---|---|---|
| HG#pRK5-EV | pRK5 | *HA* | Overexpression |
| HG#pRK5-WT | pRK5 | *ZNHIT3* | Overexpression |
| HG#pRK5-C14R | pRK5 | *ZNHIT3* | Overexpression |
| HG#pRK5-C14F | pRK5 | *ZNHIT3* | Overexpression |
| HG#pRK5-S31L | pRK5 | *ZNHIT3* | Overexpression |
| HG#pRK5-Δ251-254 | pRK5 | *ZNHIT3* | Overexpression |
| HG#FUGW | FUGW | *EGFP* | Lentiviral, Overexpression |
| HG#FUGW-WT | FUGW | *ZNHIT3* | Lentiviral, Overexpression |
| HG#FUGW-C14R | FUGW | *ZNHIT3* | Lentiviral, Overexpression |
| HG#FUGW-251-254 | FUGW | *ZNHIT3* | Lentiviral, Overexpression |

**Table S2. List of primers used in this study.**

| Primer | Sequence | Purpose |
|---|---|---|
| ZNHIT3_C14R | CGTCTGCGTGATCCGCTTGGAGAAGCC | Site-directed mutagenesis |
| ZNHIT3_Δ251-254 | GTGATGAGGAAGCAGAGTTTCTTTGC | Site-directed mutagenesis |
| ZNHIT3_S31L | GCGTGCCCTACTGCTCCGTAGTCTGCTTCC | Site-directed mutagenesis |
| ZNHIT3_C14F | CGTCTGCGTGATCTTCTTGGAGAAGCCCAA | Site-directed mutagenesis |
| ZNHIT3_BamHI_For | CACCAAGGATCCATTCGCGCCACCATGGGCTA | Cloning |
| ZNHIT3-EcoRI-Rev | CTTACTTACGAATTCGGCCGCTTAAGACTCC | Cloning |
| SNORA30-For | GCACTTTCACAGTTCCTTCC | RT-qPCR |
| SNORA30-Rev | CAGGGCAAGAATACAATCAAGG | RT-qPCR |
| SNORA55-For | AGATGGTGCTACAGAATGAGC | RT-qPCR |
| SNORA55-Rev | AAATGTGGCAGAGCTGTAGAG | RT-qPCR |
| SNORA5A-For | CAGTACCTGTCCTATGCATGG | RT-qPCR |
| SNORA5A-Rev | ACCAAATTTATCCCTGAGCCT | RT-qPCR |
| SNORD91B-For | GTCTGAACCTGTCTGAAGCATCC | RT-qPCR |
| SNORD91B-Rev | AAGCCTCAGTATCACACAGAAGT | RT-qPCR |
| SNORD93-For | GCCAAGGATGAGAACTCTAATCTGA | RT-qPCR |
| SNORD93-Rev | GGCCTCAGGTAAATCCTTTAATCCA | RT-qPCR |
| SNORD114-10-For | GATCAATGATGACTACTGTT | RT-qPCR |
| SNORD114-10-Rev | GGACCTCAGAGTTTCAGACA | RT-qPCR |
| SNORD112-For | TGGACCAATGATGAGACAGTG | RT-qPCR |
| SNORD112-Rev | GTGCAGAACTGGATTAATCATG | RT-qPCR |
| SNORD125-For | CCCTCCTGATGATTCTTCTTCC | RT-qPCR |
| SNORD125-Rev | GTCAACTTCTTAGAGGCTCAGTT | RT-qPCR |
| U3A-For | TAGCAGAGGTGTGCAAGGAGG | RT-qPCR |
| U3A-Rev | GCAGTTGCAGCCAAGCAACG | RT-qPCR |
| U14A-For | GATTGGTTGCCAGACATTCG | RT-qPCR |
| U14A-Rev | CACTCAGACATCCAAGGAAG | RT-qPCR |
| U18A-For | TAGTGATGAAATTCCACTTC | RT-qPCR |
| U18A-Rev | CATCAGAACATCCGAGAAAA | RT-qPCR |
| MRP-For | TATCCTAGGCTACACACTGAGG | RT-qPCR |
| MRP-Rev | CTTCTTGGCGGACTTTGGA | RT-qPCR |
| 7SL-For | AGGAGTTCTGGGCTGTAGT | RT-qPCR |
| 7SL-Rev | TTTGACCTGCTCCGTTTCC | RT-qPCR |
| 28S rRNA-For | AGTCGGGTTGCTTGGGAATGC | RT-qPCR |
| 28S rRNA-Rev | CCCTTACGGTACTTGTTGACT | RT-qPCR |
| 18S rRNA-For | CGGCTACCACATCCAAGG | RT-qPCR |
| 18S rRNA-Rev | TACAGGGCCTCGAAAGAGTC | RT-qPCR |
| 5.8S rRNA-For | ACTCTTAGCGGTGGATCACTC | RT-qPCR |
| 5.8S rRNA-Rev | AAGCGACGCTCAGACAGG | RT-qPCR |
| 5S rRNA-For | GGCCATACCACCCTGAACGC | RT-qPCR |
| 5S rRNA-Rev | CAGCACCCGGTATTCCCAGG | RT-qPCR |

**Table S3. Properties of the tested SNORDs.**

| Name | Host gene | ΔG (kcal/mol) | snoRNA length | Modification | Length (guide) | GC % (guide) | Tm °C (guide) |
|---|---|---|---|---|---|---|---|
| SNORD21 | RPL5 | -23.10 | 95 | 28S−Gm1303 | 15 | 46.7 | 39.2 |
| SNORD31 | SNHG1 | -17.50 | 68 | 28S−Gm4166 | 12 | 66.7 | 40 |
| SNORD72 | RPL37 | -19.20 | 80 | 28S-Um4590 | 15 | 26.7 | 31 |
| SNORD91B | TSR1 | -58.70 | 222 | 28S−Gm4588 | 16 | 37.1 | 38.3 |
| SNORD93 | SNHG26 | -21.70 | 74 | 18S−Am576 | 13 | 23.1 | 32 |
| SNORD125 | AP1B1 | -31.20 | 96 | 18S-Cm1440 | 15 | 39.5 | 36.5 |
| SNORD126 | CCNB1IP1 | -18.20 | 77 | 28S−Am1310/ 18S (predicted) | 13/8 | 45.5 | 36 |
